# High failure rate of ChAdOx1 in healthcare workers during Delta variant surge: A case for continued use of masks post-vaccination

**DOI:** 10.1101/2021.02.28.21252621

**Authors:** Rajat Ujjainiya, Akansha Tyagi, Viren Sardana, Salwa Naushin, Nitin Bhatheja, Kartik Kumar, Joydeb Barman, Satyartha Prakash, Rintu Kutum, Akash K Bhaskar, Prateek Singh, Kumardeep Chaudhary, Menka Loomba, Yukti Khanna, Chestha Walecha, Rizwan Ahmed, Ashutosh Yadav, Archana Bajaj, Gaurav Malik, Sahar Qureshi, Swati Waghdhare, Samreen Siddiqui, Kamal Krishan Trehan, Manju Mani, Rajiv Dang, Poonam Das, Pankaj Dougall, Monica Mahajan, Sudipta Sonar, Kamini Jakhar, Reema Kumar, Mahima Tiwari, Shailendra Mani, Sankar Bhattacharyya, Sandeep Budhiraja, Anurag Agrawal, Debasis Dash, Sujeet Jha, Shantanu Sengupta

## Abstract

Immunization is expected to confer protection against infection and severe disease for vaccinees, while reducing risks to unimmunized populations by inhibiting transmission. Here, based on serial serological studies, we show that during a severe SARS-CoV2 Delta-variant outbreak in Delhi, 25.3% (95% CI 16.9 - 35.2) of previously uninfected, ChAdOx1-nCoV19 double vaccinated, healthcare-workers (HCW) were infected within a period of less than two months, based on serology. Induction of anti-spike response was similar between groups with breakthrough infection (541 U/ml, IQR 374) or not (342 U/ml, IQR 497), as was induction of neutralization activity to wildtype. Most infections were unrecognized. The Delta-variant thus causes frequent unrecognized breakthrough infections in adequately immunized subjects, reducing any herd-effect of immunity, and requiring reinstatement of preventive measures such as masking.

## Main

Immunization is expected to confer protection against infection and severe disease for vaccinees, while also indirectly protecting unvaccinated populations by reducing transmission. Initial data for SARS-CoV2 vaccines suggested that break-through infections would be infrequent and be associated with lower viral loads, shorter duration, and low likelihood of transmission ^1^. Easing of social restrictions such as universal masking was critically dependent on validity of these initial observations. However, the global surge in transmission of Delta variant of SARS CoV2, including in high vaccination populations such as Israel ^2 3^, has led to reconsideration of the policies ^4^. More recent data suggests that Delta infections have higher viral loads, with no difference between vaccinated and unvaccinated ^5 6 7 8^. Together, the data strongly argues that frequent breakthrough infections and minimally impeded transmission is a possibility with new variants such as Delta.

In healthcare setting, transmission from vaccinated healthcare workers (HCW) to patients is a particular concern since many patients may be high-risk. Vaccine efficacy studies have typically used rtPCR positivity as the primary instrument for determining breakthrough rates. The true rate of infection is likely to be much higher since vaccination is known to reduce symptoms and increase test-seeking threshold. Serologic indicators of infection can overcome these limitations, with the caveat that we do not fully know about the transmissibility of asymptomatic infections discovered by serologic testing. However, epidemiologic models have clearly shown that such infections contribute substantially to aggregate community transmission ^9^.

Recently, there was a severe Delta-variant outbreak in Delhi, India ^8^. We have shown that the odds of vaccination breakthroughs were greatly increased by Delta-variant, formed larger clusters than seen previously, and were associated with high viral loads ^7^. We also found that there was very high community transmission, with seropositivity in a small cohort rising from 42% before the outbreak to 86% afterwards ^8^. However, the first study in vaccinated HCW lacked serological assessments, while the latter was in an unvaccinated population. Here, we provide bridging data for serological estimates of vaccination breakthroughs in a HCW cohort, where the Delta outbreak was coincidentally bracketed between serial sample collection time-points (Fig 1A). The structure of the cohort and data available for each time-point is shown in figure 1B.

**Figure 1:**
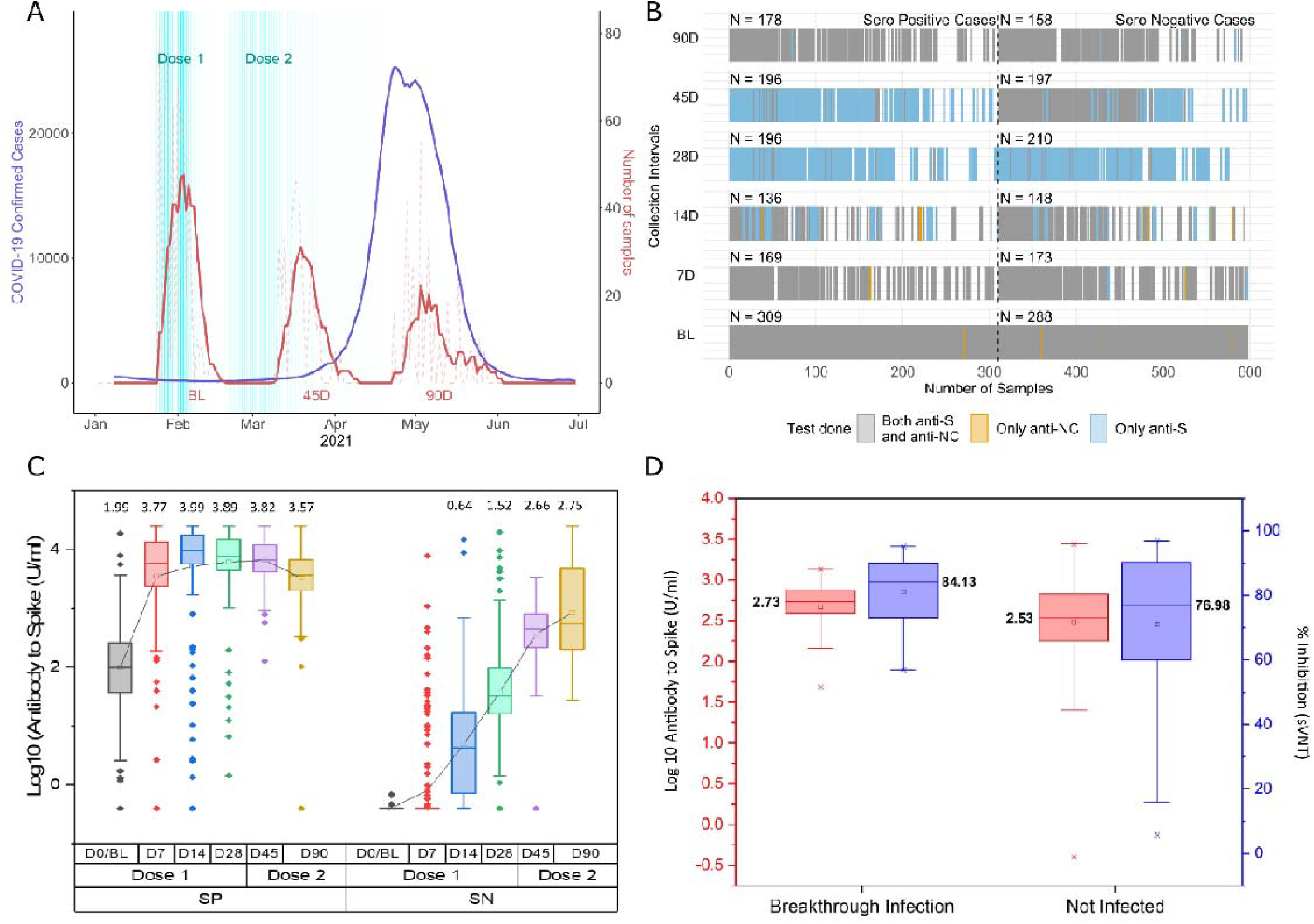
A) Time distribution of sampling for HCW cohort shown alongside the Delta-variant driven surge (April, May 2021). Sample collection is 7 days moving average for HCW cohort. B) Data Structure of the cohort and sampling C) Antibody to spike-protein in log_10_ units (U/ml) at baseline (D0/BL) and 7, 14, 28,45, 90 days after first dose in baseline antibody naïve (SN) and infection recovered (SP) subjects. Second dose precedes D_45_ and D_90_. D) Antibody to Spike in log_10_ units (U/ml) and % Inhibition (sVNT) response for breakthrough cases and non-infected subjects after two doses of vaccine in D_0_(Baseline/BL) antibody naïve individuals.

Briefly, the cohort contained 597 ChAdOx1-nCoV19 vaccine recipients. The timing between first and second dose varied, but 482 received the second dose within 42 days of the first dose and most subjects received the second dose at 28±3 days. Fifty-two percent subjects (n=309) had been previously infected with SARS-CoV-2, based on presence of antibodies to SARS-CoV2 proteins (spike, anti-S or nucleocapsid, anti-NC) at D_0_, the day of first dose. There was a robust immunogenic response to two-doses of vaccine, irrespective of prior infection (Figure 1C). A single dose seemed sufficient for previously infected individuals in terms of absolute antibody levels as well as neutralization activity (sVNT) (Figure 1C, and Supplementary Fig 1A).

Since ChAdOx1 immunization only induces Anti-S, subsequent anti-NC seroconversion in previously anti-NC negative subjects was taken as a sensitive and specific marker of new infection^10-12^. A sharp increase in Anti-NC concentration after a decline can be taken as a specific marker of reinfection, but the pattern requires a minimum of three consecutive samples and sufficient time for decline, making it prone to false negatives^8^. Our primary focus was thus on quantifying new breakthrough infections in vaccinated subjects, defined as [Anti-NC^negative^ at D_45_ AND Anti-NC^positive^ on D_90_. We also estimated breakthrough reinfections using the conservative decline and rise definition above, as well as a more relaxed criteria based on doubling of anti-NC (see supplement).

Amongst fully vaccinated and uninfected HCW, i.e. completed 2 weeks beyond the second dose and Anti-NC^negative^ at D_45_, the breakthrough infection prevalence at D_90_ was 25.3% (95% CI 16.9 - 35.2). Comparison of Anti-S concentration and sVNT between D_45_ sera of the breakthrough infection group and those stayed uninfected (Anti-NC^negative^ at D_45_ and D_90_) is shown in Figure 1D. Data for those HCW who were previously infected had a reinfection rate of 2.5% over the same period. Through follow up of a general employee cohort, where most subjects had received only a single dose of ChAdOx1, we observed that 48.4% (95% CI 30.2 - 66.9) of previously uninfected subjects had a vaccination breakthrough after single dose, within the period of this study. The questionnaire survey indicated that there were no severe infections leading to hospitalizations in either of the two cohorts.

The data indicates that ChAdOx1-nCoV19 immunization in our cohort yielded about the same degree of humoral immune response as has been reported elsewhere in terms of Anti-S levels ^13-15^. This would indicate generalizability of our observations to other populations. Further, Anti-S levels correlated well with MnT assay to both WT and Delta. However, neutralization activity for equivalent levels of Anti-S was much lower for Delta than WT, and conversely higher Anti-S levels were required for comparable neutralization of Delta (Figure 2A). Pre-infection anti-S level greater than 1500 U/ml was required for neutralization titres of 1:80 or better against the Delta variant. Such levels were not commonly seen after vaccination unless there has been previous infection. In our data, 2.4% (3 of 125) subjects who were Anti-NC^negative^ at D_0_ and D_45_ had Anti-S levels higher than 1500 U/ml at D_45_, compared to 96.4% (162 of 168) of subjects who were Anti-NC^positive^ at D_0_. We noted excellent post-breakthrough infection neutralization activity to Delta, consistent with likely Delta-variant related breakthroughs and rise in Anti-S and Anti-NC levels (Figure 2B and 2C).

**Figure 2:**
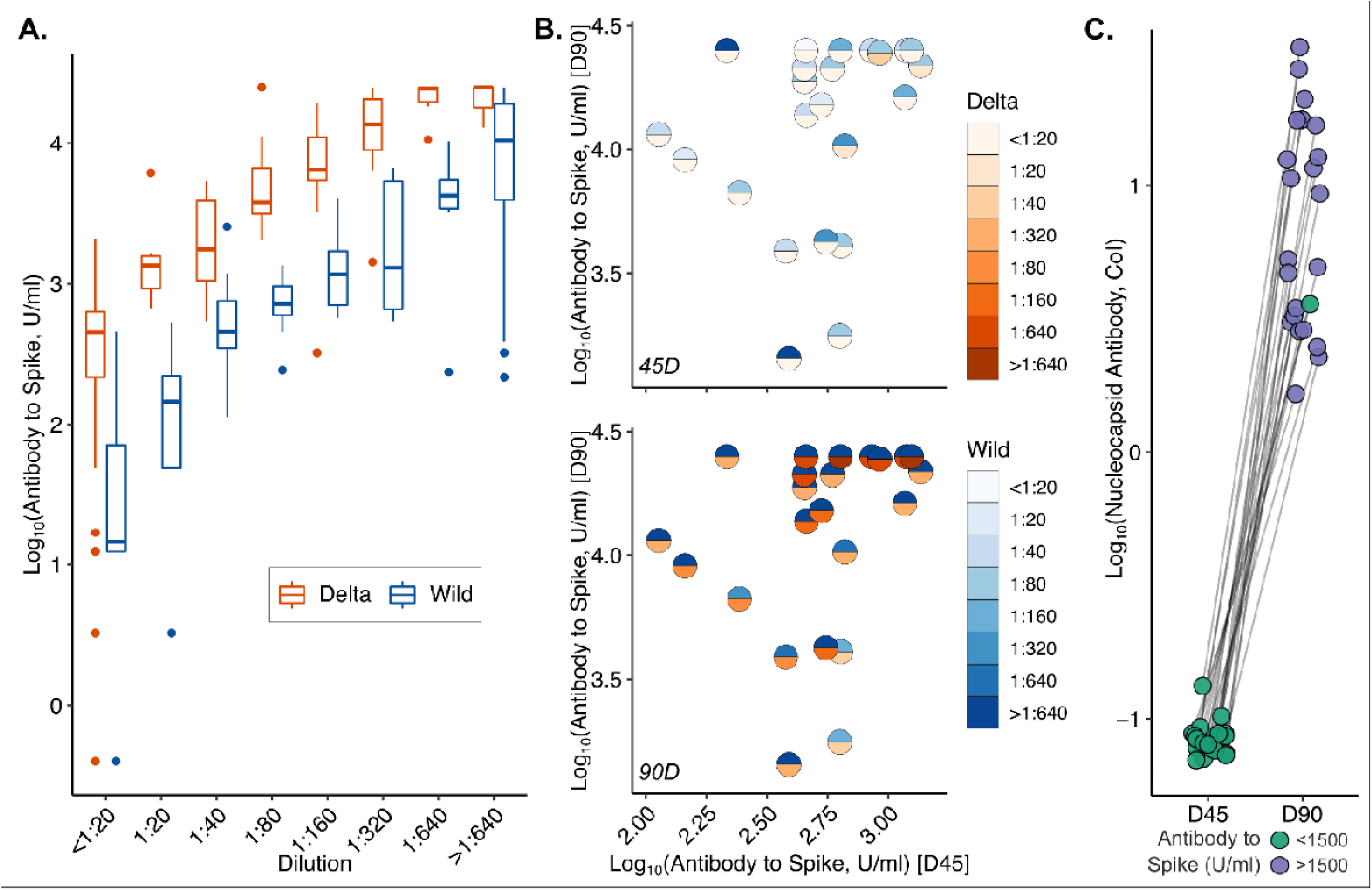
A) Box plot for Anti-S titres for similar neutralization titres against Wild type and Delta strain. (B) MnT assay titre to the Wild strain and Delta strain response at day 45 and 90 for breakthrough cases C) Anti-NC antibody response at D_45_ and D_90_ when comparing rise in quantitative titres for breakthrough cases.

The primary limitations of our data are that we did not have the necessary information to determine whether there were transmission chains with breakthrough infections, nor have we confirmed that these were Delta infections. While we have identified transmission clusters in other published data ^7^, the two situations are not directly comparable, with one being a serological diagnosis and the other being RT-PCR. Given the known relationships between RT-PCR positivity and humoral response, where seroconversion occurs in a subset of RT-PCR positive subjects, it seems reasonable to assume that infection sufficient to induce humoral response would also have been RT-PCR positive at some point ^16^.

Within these limitations, our results have three important implications for management of Delta outbreaks, some of which have already been confirmed by other data. First, neutralization of Delta variant by antibodies to non-Delta spike protein is greatly reduced (Figure 2). This means that neither prior infection by non-Delta, nor existing vaccines, are individually sufficient for the path to herd immunity. This also implies that masking is an essential part of any rational COVID control strategy, being agnostic to immune escape. Second, given reduced effectiveness of induced antibodies, a single dose of ChAdOx1-nCoV19 should not be expected to confer any protection against Delta variant infection, and second dose should be given early, preferably within 6 weeks, as was done for HCW here. We found that 48.4% (30.2 - 66.9) of previously uninfected subjects had a vaccination breakthrough after single dose, within the period of this study. That is unacceptably high.

Last, our data indicates an urgency to explore routes towards more effective use of vaccines. Even one dose of ChAdOx1-nCoV19 to previously infected subjects induces humoral immunity comparable or better than two doses in naïve subjects. This can be a preferred strategy for optimal use of vaccines in populations with high seropositivity. In susceptible populations, there may be benefit to using heterologous prime/boost strategies as shown by the ComCov study ^13 17^, where using ChAdOx1-nCoV19 as prime followed by BNT162b2 mRNA boost provided statistically higher levels of humoral immunity comparable to homologous ChAdOx1 (12906 ELU/ml vs 1392 ELU/ml). Cellular immunity may be better by this approach and the decline may be slower, although this remains to be verified.

To conclude, our data shows that vaccination breakthroughs were far more common during the Delta-outbreak in Delhi than previously reported. Protection against Delta during high exposure is only reached at very high levels of ChAdOX1-nCoV19 induced Anti-S and prior infection was seen in most subjects who crossed this threshold. Natural infection was seen to provide a strongly boosted response to vaccination. There is a need to have a variant-specific high throughput neutralization assay to truly assess population immunity and predict outbreak risk.

## Methods

The study was approved by the ethics committees of Max Group of Hospitals, New Delhi and CSIR-IGIB, New Delhi. Healthcare workers (HCW’s) at the Max Hospital Group, who were to receive the ChAdOx1-nCoV19 vaccine, voluntarily enrolled after an informed consent. Quantitative antibody response directed against the spike protein was measured at day 0, 7, 14, 28, 45 and 90, with qualitative antibody response on day 0,45 and 90 and neutralizing antibody response on day 28, 45 and 90. At day 45, we assessed subjects who had received their second dose at 28±7 days and provided their sample at day 45±3 days. Similarly, for data at 90 days we included who got their second dose up to 42 days and gave their sample at 90±20 days. Herein, we analysed data of 597 ChAdOx1-nCoV19 vaccine recipients. The timing between first and second dose varied, but 482 received the second dose within 42 days of the first dose and most subjects received the second dose at 28±3 days. Of these; 309 (52%) had already developed antibodies to SARS-CoV-2 HCW’s were thus divided into two groups - seropositive (SP) and seronegative (SN), based on their serology (Anti-NC) status at baseline. We were also following a second CSIR cohort of which first phase had been completed ^18^ with Phase 2 data collected during Jan-Feb 2021, the third phase collection was ongoing between May to July 2021. ^8^ Two separate cohorts were made for assessment of vaccination response and breakthrough for the HCW’s as frontline workers had to necessarily receive the two doses at 28 day dosing interval in the initial period of vaccination program while the normal population from March-April onwards had delayed dosing interval and hence the cohort was amenable to monitoring for vaccination response against breakthrough infection when only a single dose was taken and also comprising of individuals who had not taken any vaccine to the date of sample collection and thus could be assessed for natural infection assessed both from history and serological response through nucleocapsid antigen based qualitative antibody response.

### Sample Collection

Blood samples (6 ml) were collected in K2 EDTA vials from each participant and antibodies to SARS-CoV-2 directed against the spike protein (S-antigen) were assayed using Elecsys Anti-SARS-CoV-2 S quantitative antibody detection kit (Roche Diagnostics), according to the manufacturer’s protocol. Antibody levels >0.8 U/ml considered were seropositive. The range of the kit is from 0.4 U/ml to 250 U/ml. For samples with values of >250 U/ml appropriate dilutions were made. Samples were also assessed for qualitative antibody (Anti-NC) using the same manufacturer’s kit. Samples on Day 28, 45 and 90 were further tested for neutralizing antibody (NAB) response directed against the spike protein using GENScript cPass™ SARS-CoV-2 Neutralization Antibody Detection Kit (Genscript, USA), according to the manufacturer’s protocol.^18^

In this report we have performed microneutralization assay using live virus. For this purpose SARS-CoV-2 virus belonging to the original Wuhan type strain and Delta strain were grown in VeroE6 cells and the infectious titre determined by Reed and Muench method. Aliquots of the virus were stored in a -80 freezer inside BSL3 laboratory in single use aliquots. Serum samples were heat inactivated at 56°C for 1 hour and subsequently stored at -80 freezer till assay. 10,000 VeroE6 cells from an exponentially growing culture were seeded per well in a 96-well plate for at least 20 hours before the assay. On the day of assay, 2-fold serial dilutions of the serum were prepared in serum-free media (SFM) and the plate shifted to BSL3. Each serum dilution was mixed with 100 TCID50 of SARS-CoV-2 virus and incubated at 37°C, 5% CO_2_ for 1 hour. The mix of virus and serum dilutions were then allowed to infect the VeroE6 cell monolayer in the 96-well plate, for 1 hour at 37°C, 5% CO_2_. The inoculum was discarded and growth media supplemented with 2% foetal calf serum overlaid on the monolayers. The plates were incubated at 37°C, 5% CO_2_ for 72 hours. The cytopathic effect from the virus infection was scored visually using a phase-contrast microscope. The controls used included a positive control (no CPE) and a negative control serum (visible CPE) in addition to a mock-infection control (no CPE). The highest dilution of an experimental serum that inhibited appearance of visible CPE was scored as the neutralization titre corresponding to that serum for the strain of virus used.

### Statistical Analysis

Data analysis was carried out with visualization in MS-Excel 2016, OriginPro V2021 and R version 4.0.3. A non-Parametric Mann-Whitney test was utilized for SN and SP group comparison, for time specific significant differences.

## Data Availability

Requests for de-identified data may be send to the corresponding authors stating end usage. Data will be shared on request basis.

## Author Contribution

SSG, SJ, DD and AA conceptualized the idea and coordinated the research activity. RU, SN, SSG, AKB, KK, JB contributed in the sample analysis, storage and sVNT based neutralization antibody experiments. DD, VS, SSG, AA, NB, AT, SPrakash and RU contributed in Data Acquisition, Data Pre-Processing, Data Analysis. VS, SSG, DD, AA, RK and PS contributed in visualization. AT, ML, YK, CW, RA, AY, AB, GM,SQ, SW, SS, KKT, MM, RD, PD, PDougall, MMahajan, SBudhiraja contributed in HCW centre co-ordination, enrolment of volunteers, sample collection and logistics. SSonar, KJ, RKumar, MT, SM, SB contributed in MnT based assay and analysis. VS and SSG wrote the first draft. AA, SSG, VS, DD and KC revised the manuscript. All the authors agreed and contributed to the final draft of the manuscript.

## Acknowledgment

SSG would like to acknowledge CSIR grant MLP 2007 for this work. The following reagent was deposited by the Centers for Disease Control and Prevention and obtained through BEI Resources, NIAID, NIH: SARS-Related Coronavirus 2, Isolate USA-WA1/2020, NR-52281. SSG would like to acknowledge Manish Chowdhary, Yasmeen Khan and Preeti Subramani for sample analysis. SJ, SSG and DD would like to acknowledge all participants of the study.

