## Supplement for "High failure rate of ChAdOx1 in healthcare workers during Delta variant surge: A case for continued use of masks post-vaccination"

*Immunogenic Response with two doses of vaccine*

Antibody response at 7 days in the Seropositive (SP) group was significantly higher than in the Seronegative (SN) group (p<0.0001) In fact, greater than ninety percent of subjects in the SN group did not develop any measurable response at day 7. This was corroborated at day 14 when subjects in the SN group started developing antibodies, though the difference was still significant amongst the two groups (p<0.0001). On day 28, before the second dose of vaccination, most of the individuals in the SN group had measurable antibody response. (Figure 1A)

At day 45, we assessed subjects who had received their second dose at 28±7 days and provided their sample at day 45±3 days. Similarly, for data at 90 days we included who got their second dose up to 42 days and gave their sample at 90±20 days. Interestingly, in the SP group, there was no further increase in the antibody levels after administration of the second dose and were observed to fall at day 90. However, in the SN group, the antibody levels kept rising after the second dose at day 45 but did not reach the levels of baseline seropositive group albeit at D_90_ the levels were stable in comparison to day 45 and were not observed to be falling unlike the SP group where nearly a two-fold decline was observed (Figure 1A).

Median levels for nAb (sVNT), which had already peaked at D_28_ remained stable at 97% in the SP group after the second dose when observed at day 45 and day 90. On the contrary, in the SN group the median level of the neutralizing antibody (sVNT) was 44% after the first dose which increased to 81% after the second dose. (Supplementary Figure 1).

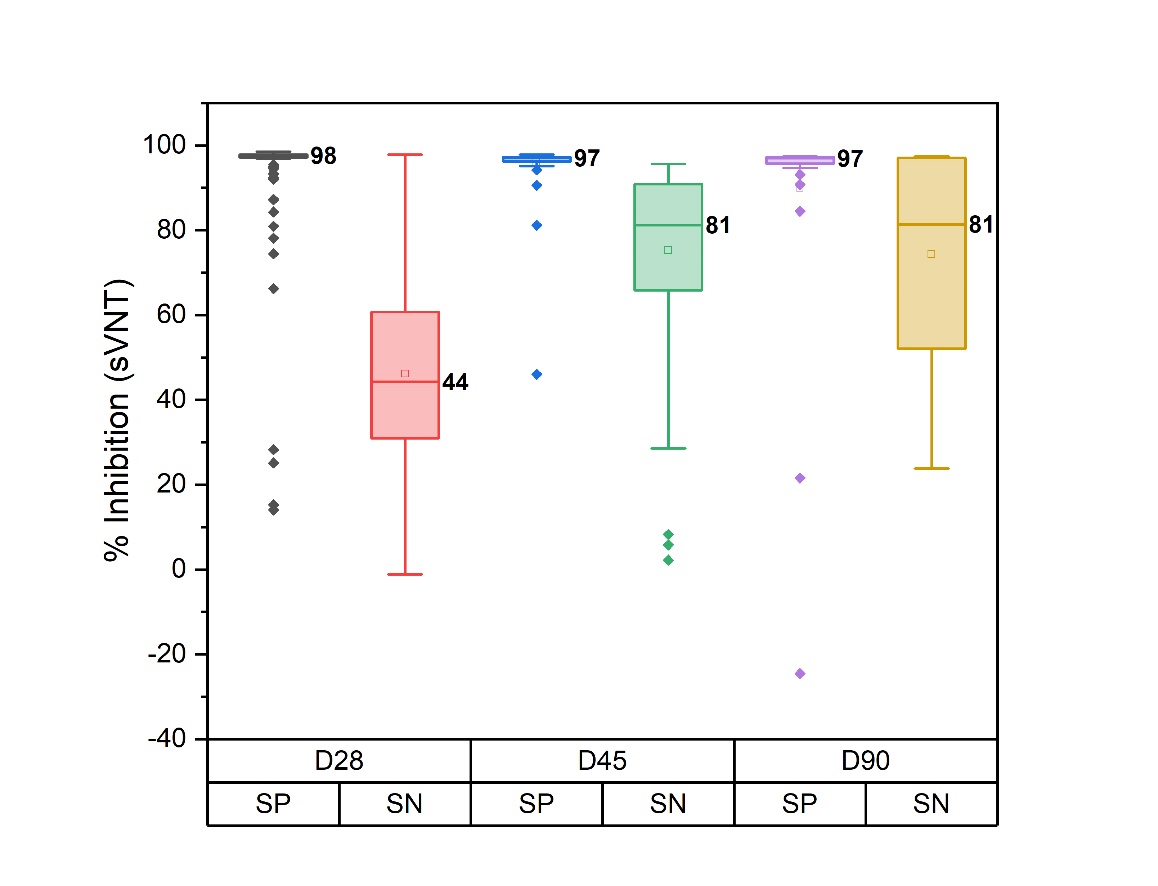

Supplementary Figure 1: nAb response (sVNT) assay at D_28_, D_45_ and D_90_ in SP and SN subjects (Values are rounded off).

*Relaxed Criteria*

To assess breakthrough, the criteria was;

Setting the dosing interval up to D_42_, the subject should be Anti-NC negative at D_45_ and when followed to D_90_ should show a positive CoI i.e. CoI>=1. (D90 follow up included samples collected D_70_ onwards from 1^st^ dose

However, under relaxed criteria, at D_90_ the CoI at D_90_ could be between 0.1 and 1, but, should show Anti-NC increase greater than two-fold and Anti-S increase greater than five-fold to qualify as a breakthrough.
